# Explaining the Variance in Cardiovascular Health Indicators among Asian Americans: A Comparison of Demographic, Socioeconomic, and Ethnicity

**DOI:** 10.1101/2024.07.30.24311252

**Authors:** Lalaine Sevillano, Adrian Matias Bacong, Dale Dagar Maglalang

**Author notes:** **Corresponding author:** Lalaine Sevillano, School of Social Work, Portland State University, 1800 SW 6th Avenue, Portland, OR, 97219.

## Abstract

**BACKGROUND:** The Asian American (AA) population is the fastest-growing major racial or ethnic group in the U.S. Typically treated as a monolith in research, disaggregated data show disproportionate cardiovascular disease (CVD) burden among certain AA ethnic groups. This analysis aimed to identify which factors explain variance in cardiovascular health among AA ethnic groups.

**METHODS:** We analyzed pooled 2010-2018 National Health Interview Survey cross-sectional data from Chinese, Asian Indian, Filipino, and Other Asian adults in the U.S. Coronary heart disease, heart attack, and stroke were the CVD outcomes of interest. Covariates included sociodemographic characteristics, CVD-related health behaviors (e.g., smoking tobacco, physical inactivity), and health conditions (e.g., diabetes, hypertension). The distribution of self-reported CVD outcomes and covariates were examined among the full AA sample and disaggregated ethnic groups. Variance explained by sociodemographic, health behaviors, and health conditions were calculated based on the adjusted R-squared from a series of five models for each CVD health outcome.

**RESULTS:** Of the 10,353 AAs in the sample, 53% identified as female and 86% between the ages of 18-64 years old. Compared to the aggregate AA sample and the other ethnic groups, Filipinos had a higher burden of any CVD outcome (5.9%), particularly for coronary heart disease (4.0%) and heart attack (2.5%).The combination of all predictors explained at most 13% of variance, with sociodemographic characteristics accounting for at least half of the variance explained among all participants. Health behaviors explained a greater amount of additional variance for all CVD outcomes among Asian Indians, including an additional 3.1% for stroke. Inversely, existing health conditions were significant predictors of CVD for all AA ethnic groups compared to Asian Indians.

**CONCLUSIONS:** There is heterogeneity in CVD outcomes and related risk factors in AA ethnic groups, emphasizing the need for culturally-tailored prevention and intervention strategies.

## INTRODUCTION

The Asian American (AA) population currently constitutes 7% of the total United States (U.S.) population, nearly doubling its size from 11.9 million back in 2000 to 22 million today.^1^ The AA population is the fastest-growing major racial or ethnic group, with a growth rate of nearly four times faster than the overall U.S. population.^2^ By 2060, AAs are estimated to have a population of 46 million.^1^ Among the primary contributors to mortality in the AA population is cardiovascular disease (CVD).^3^ Despite this problem, CVD research in AAs has declined during the last decade.^3^ Additionally, despite having ancestral roots stemming from over 20 countries, AAs are often aggregated in biomedical and public health research, masking various health disparities between and within AA ethnic groups.^4^

The dearth of studies with disaggregated data indicates differences in CVD outcomes, CVD-related risk factors, and premature mortality due to CVD among AA subgroups.^4–7^ Another study using disaggregated data shows that obesity in the AA population varies; adjusting for a Body Mass Index (BMI) cut-off of 27.5 kg/m^2^ or greater, was 22.4% and highest among Filipino (28.7%), Japanese (26.7%) and Asian Indian (22.4%) and lowest among Chinese (13.2%).^8^ Other research shows that AA subgroups, except for Chinese Americans, show a higher risk for total and premature hypertension than non-Hispanic white (NHW) Americans.^9^ A comprehensive review of sleep studies on AAs determined that AAs have lower sleep duration than their NHW counterparts, but comparable sleep duration with non-Hispanic Black (NHB) groups.^10^ Within the AA population, East Asians showed better sleep outcomes than Southeast Asians.^10^ Whereas smoking has decreased in the U.S. in the past decade, national smoking prevalence in the AA population is 9.4% compared to 34% of American Indian/Alaska Native and 31.9% of multi-racial groups.^11^ However, when disaggregating data by Asian ethnic groups, cigarette use is highest in Japanese Americans (19.4%), Vietnamese Americans (18.9%), and Korean Americans (18.8%).^12^ Most recently, Lim and colleagues (2024)^13^ found significant differences in the prevalence of CVD risk factors among AA subgroups by ethnicity and birthplace. Specifically, they found that foreign-born Filipinos had a higher prevalence of all five CVD-risk factors except smoking when compared with NHW adults. Additionally, foreign-born Asian Indians had a significantly higher prevalence of diabetes, physical inactivity, and overweight/obesity, while foreign-born Chinese had a higher prevalence of physical inactivity. Collectively, these results demonstrate that CVH research with AA populations is nuanced and needs continued examination. Studies such as these underscore the importance of research that disaggregates AA ethnic groups, especially since culturally-tailored interventions have been shown to improve health outcomes for AA populations.^14–15^

Drawing from the National Institute of Minority Health Disparities Research Framework,^16^ the present study aimed to investigate heterogeneity in CVD outcomes and CVD risk factors among AAs living in the U.S. using the 2010 to 2018 National Health Interview Survey (NHIS). This ever-evolving framework highlights health disparities’ complex, multi-dimensional, and multilevel nature. Additionally, it provides a systematic approach to understanding how these determinants of health intersect and operate across the lifespan. In the current study, we locate multiple behavioral and sociocultural factors at the individual level associated with increased risk for experiencing CVD outcomes of coronary heart disease (CHD), stroke, heart attack, and any CVD. Specifically, the present study aimed to answer the following research question: Are there differences in CVD burden among AA ethnic groups (Chinese, Asian Indian, Filipino, Other Asian)?

## METHODS

### Data

We used secondary data from the 2013-2018 National Health Interview Survey (NHIS) obtained through the Integrated Public Use Microdata Series (IPUMS). The NHIS is a nationally representative health survey and a primary federal source of health information. Detailed information on the survey design and methods can be found elsewhere (http://www.cdc.gov/nchs/nhis.htm). All variables in NHIS are self-reported. The current analysis included adult respondents (aged ≥18 years) with available self-report race and/or ethnicity data. Although more recent years of data are available, the NHIS does not provide disaggregated Asian race data after 2018.

### Measures

#### Outcomes

We examined four potential outcomes of interest: reported presence of coronary heart disease (CHD), heart attack, stroke, and any CVD. Reported presence of CHD, heart attack, and stroke were asked as the following: “Have you EVER been told by a doctor or other health professional that you had CHD/heart attack/stroke?” We coded individuals who indicated “yes” as “1” and individuals who stated “no” as “0”. Individuals who refused to respond to the question or indicated “Did not know” were coded as missing. From these three outcomes, we created an “any CVD” outcome where “1” indicates a person who reported having CHD, heart attack, or stroke while “0” indicates a person who did not report having CHD, heart attack, or stroke.

#### Independent Variable

Participant ethnicity was our main independent variable. All respondents self-reported their race as one of Asian ethnicity among Chinese, Asian Indian, Filipino and ‘other Asian’ were included. Due to NHIS regulations on confidentiality, disaggregated data on ‘Other Asian’ group is not publicly available.

#### Covariates

We examined four sets of covariates as possible explanations behind disparities in CVD among each Asian group: demographic characteristics, socioeconomic factors, CVD-related health behaviors, and CVD-related health conditions. Our demographic characteristics included age in years and binary gender (male or female). Our socioeconomic factors included level of education (less than high school graduate, high school graduate or GED equivalent, some college/Associate’s or Technical Degree, college graduate or above, and missing education) and the ratio that participants’ annual income was above the Federal Poverty Level (FPL): Less than 1.00 FPL, 1.00 to 1.99 FPL, 2.00+ FPL, and Undefinable or Unknown. We included missing categories for both education and income given that these variables have high missingness in survey data. Our health behavior covariates included obesity, smoking status, hours of sleep, psychological distress, and physical activity. Body mass index (BMI) was calculated based on height and weight and obesity status defined as BMI greater or equal to 30.0 kg/m2. Tobacco use was present if participants self-reported as having ever smoked tobacco, “Have you smoked at least 100 cigarettes in your ENTIRE LIFE?” Current and former tobacco use were assessed by, “Do you NOW smoke cigarettes every day, some days or not at all?”. We coded smoking status as individuals who “Never Smoked”, “Formerly/Has Smoked”, and “Currently smokes”. The hours of sleep covariate were based on question about sleep duration categorized as “Less than 7 hours”, “7 to 9 hours”, and “More than 9 hours” based on current recommendations for adults. Finally, psychological distress was operationalized by the Kessler Psychological Distress Scale-6 (K6).^17^ We coded individuals within the following 3 groups: “No significant psychological distress (K6 < 13), “Significant Psychological Distress (K6 13+), and Missing. Finally, physical activity was determined using the International Physical Activity^18^ and coded as: “Inactive”, “Insufficiently Active”, “Sufficiently Active”, and “Missing”. Finally, our health condition covariates included the presence of hypertension and diabetes. We defined hypertension as the presence of based on the question, “Have you EVER been told by a doctor or other health professional that you had hypertension, also called high blood pressure?” Diabetes was defined as the presence of based on the question, “Other than during pregnancy, have you EVER been told by a doctor or other health professional that you have diabetes or sugar diabetes?”.

### Statistical Analysis

We first examined the distribution of the health outcomes, sociodemographic characteristics, health behaviors, and comorbid health conditions among the full sample and by disaggregated Asian groups. To determine the amount of variance explained by demographic, socioeconomic, health behaviors, and health conditions, we calculated the adjusted R-squared from a series of five models for each health outcome. Model 1 served as our baseline model, and examined the contribution of demographic factors (e.g., age and gender) on health. Our remaining models examined the contribution of our socioeconomic factors, health behaviors, and health conditions alongside the contribution of demographic factors, similar to previous work by Hamad et al. (2022).^19^ Model 2 included socioeconomic factors (e.g., educational attainment and poverty) alongside demographic factors. Model 3 included health behaviors (e.g., smoking, obesity, hours of sleep, psychological distress, and physical activity) with demographic factors. Model 4 included health conditions (e.g., diabetes, hypertension) and demographic factors. Finally, Model 5 served as our fully adjusted model which includes demographic factors, socioeconomic factors, health behaviors, and health conditions.

Similar to Hamad et al. (2022),^19^ we determined the explained variance by calculating an adjusted R-squared. Although our outcomes are binary, thus necessitating the use of a logistic modeling framework, previous work has emphasized that using ordinary least squares regression is an appropriate method to calculate the adjusted R-squared for binary outcomes.^19–22^ Given our models, we were able to examine the additional contribution of each set of covariates (e.g., socioeconomic, health behavior, health condition) above and beyond the contribution of demographic factors alone by subtracting the explained variance of our model with demographic factors only (e.g., Model 1) from our models containing additional covariates (e.g., Models 2-5), similar to Hamad et al. 2022.^19^ We also calculated the 95% confidence intervals for the adjusted R-squared for each model using the Fisher’s z-transformation and approximating the standard error for the correlation coefficient, “r”.^19, 22^ Finally, we examined each series of models separately for the full AA sample alone and by disaggregated race group. Given that models are stratified by race in disaggregated samples, adjusted R-squared values calculated are representative for that group alone (specific R-squared values and 95% CI are shown in Tables 2-6). All analyses were weighted to be representative of the aggregated AA population and the respective subgroups. We used Stata Version 17.0 for all cleaning and analysis.^23^

## RESULTS

### Sample Characteristics

The total analytical sample included 10,353 AA participants with about 53% female and 86% between the ages of 18-64 years old (Table 1). Disaggregated by ethnicity, Asian Indians were more likely to be male (51%) and younger with 52% being between 18-39 years of age. Socioeconomically, 53% of all AA participants had at least a Bachelor’s degree and 69% had a +2 ratio of annual income to FPL. Education attainment varied across ethnic groups with 72% of Asian Indians having at least a Bachelor’s degree compared to only 42% of Filipino participants. Additionally, 76% of Asian Indians had a +2 annual income to FPL ratio compared to only 62% of Other Asian participants.

**Table 1.**
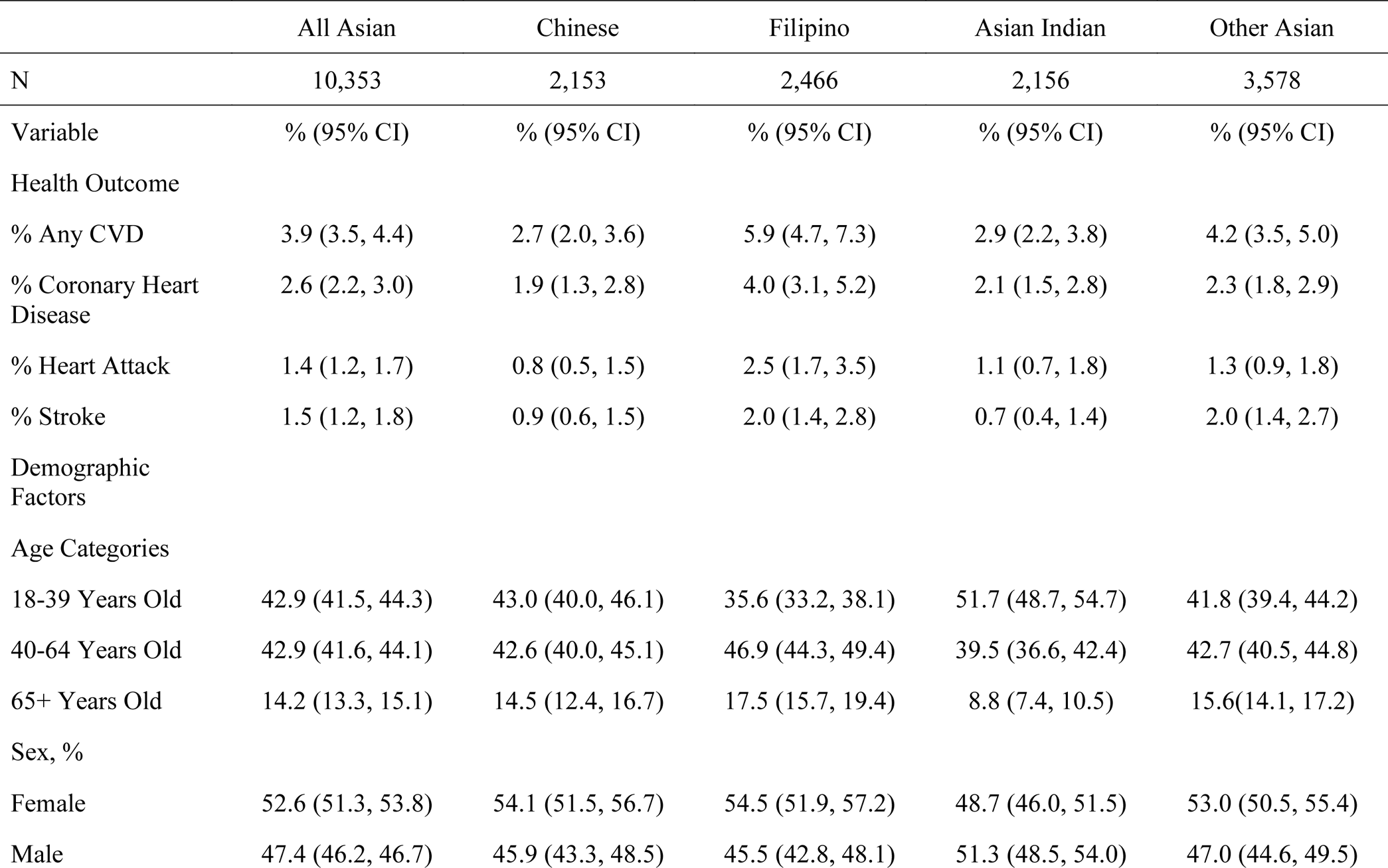
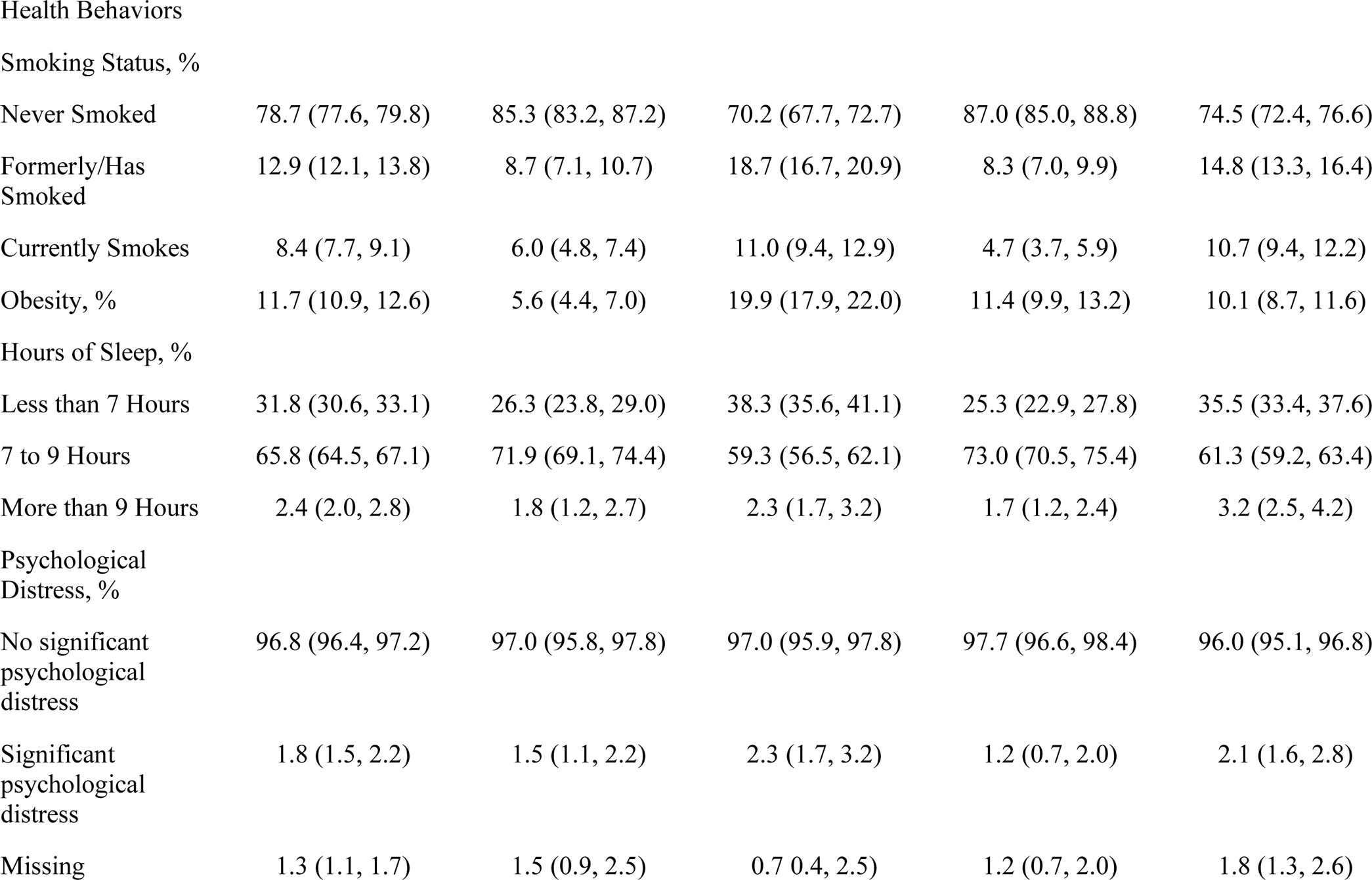
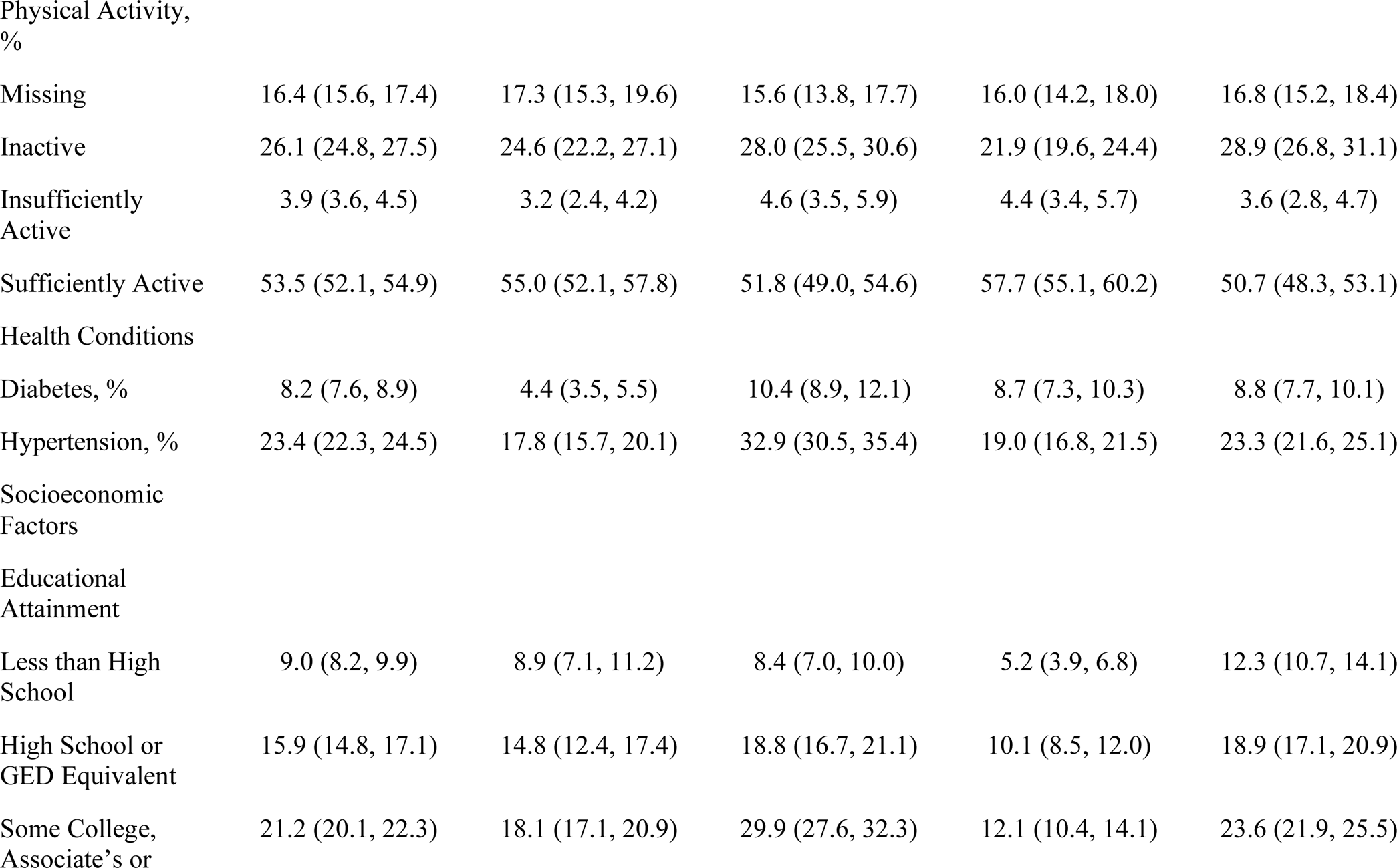
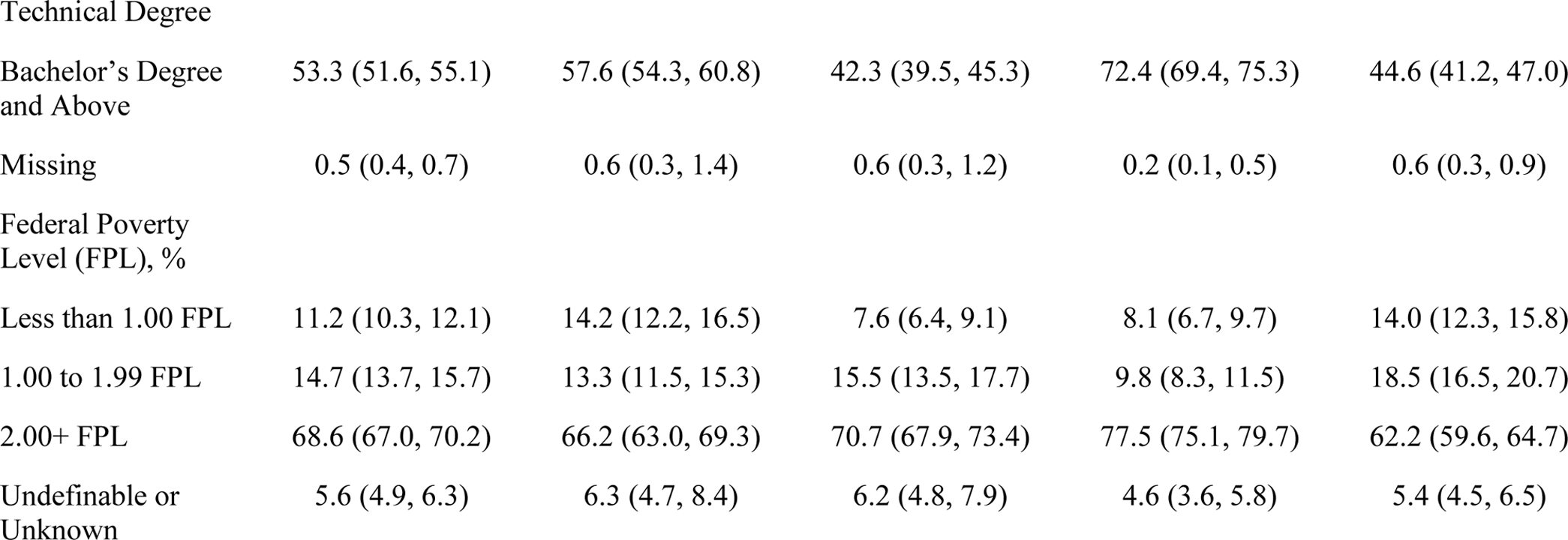
Weighted Participant Characteristics by Disaggregated Asian Ethnic Group, N = 10,353.

### Variance in CVD Explained by Demographic Characteristics

R-squared values for regression models incorporating the demographic characteristics of age and gender (blue bars in Figure 1, Panels A-E) ranged from 2.7% heart attack to 7.0% any CVD for all AA participants; 1.5% stroke to 4.5% any CVD among Chinese participants (Figure 1, Panel B), from 3.8% stroke to 8.7% any CVD among Filipino participants (Figure 1, Panel C), from 2.2% stroke to 8.0% any CVD among Asian Indian participants (Figure 1, Panel D), and from 1.7% heart attack to 6.6% any CVD among Other Asian participants (Figure 1, Panel E). The total variance explained for all categories of predictors was below 14.0% for all outcomes, ranging from 4.9% heart attack to 11.0% for any CVD among all AA participants (Table 2), 5.5% heart attack and 8.8% for any CVD among Chinese participants (Table 3), 6.1% stroke to 13.1% for any CVD among Filipino participants (Table 4), 4.9% heart attack to 12.6% any CVD for Asian Indian participants (Table 5), and 6.3% heart attack to 12.5% any CVD for Other Asian participants (Table 6). For CHD, heart attack, stroke, and any CVD, demographic characteristics accounted for at least half of the variance that could be explained by all predictors combined among all participants. However, when data were disaggregated by ethnic group, demographic characteristics accounted for at least half of the variance explained for all outcomes only among Filipino participants. Among Chinese, Asian Indian, and Other Asian participants, demographic characteristics accounted for at least half of variance that could be explained by all combined predictors for only any CVD and CHD.

**Figure 1.**
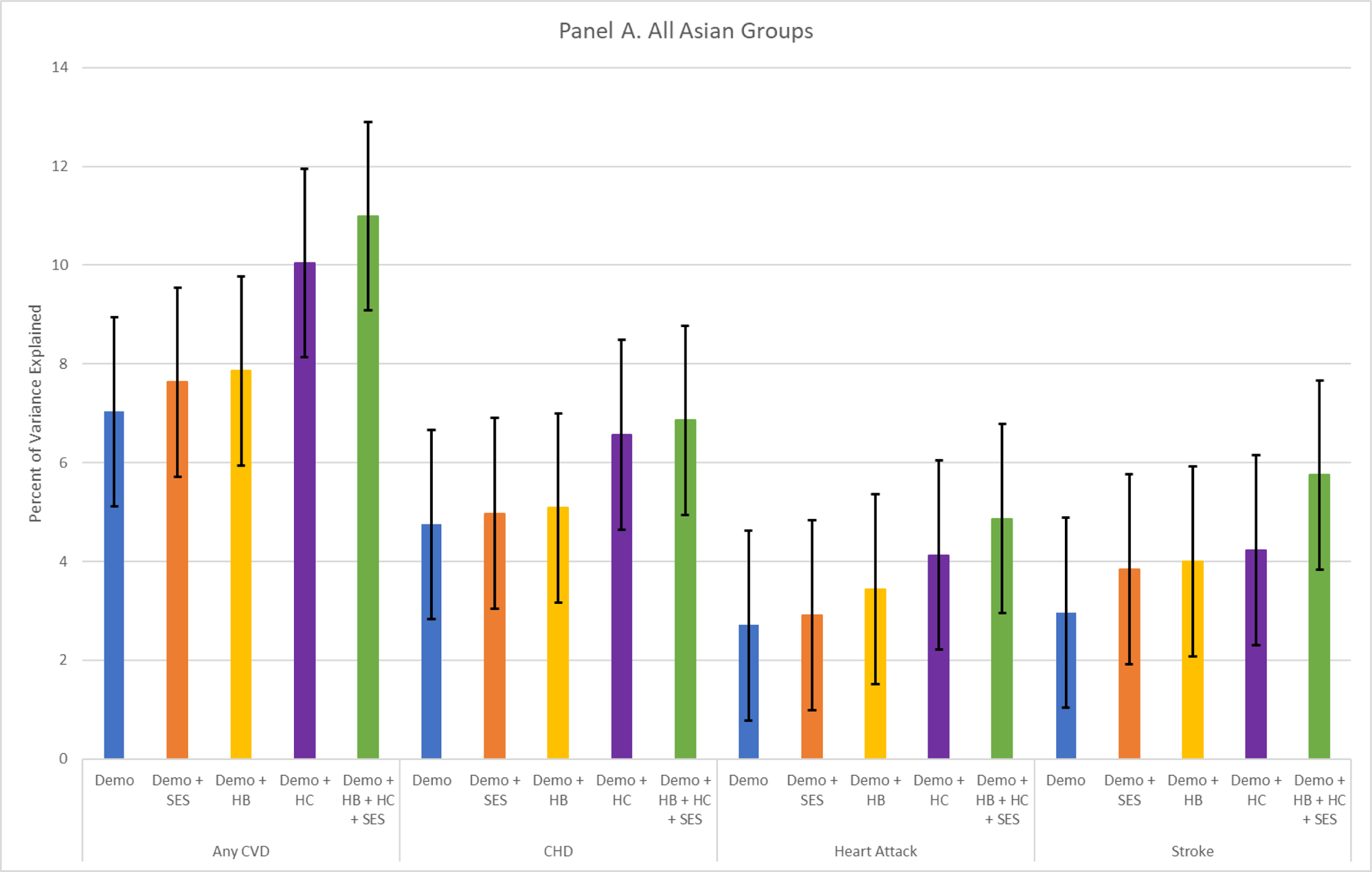
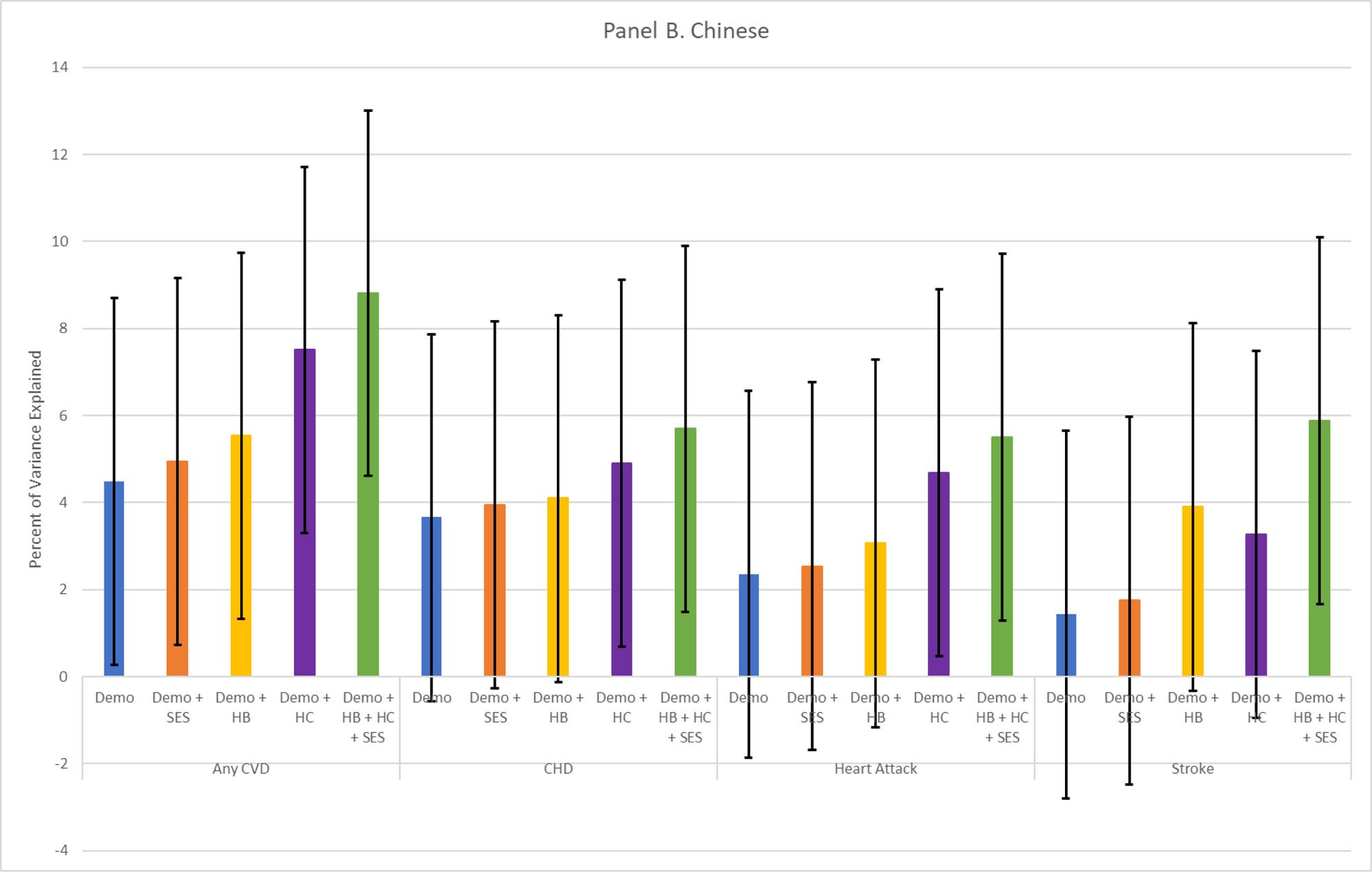
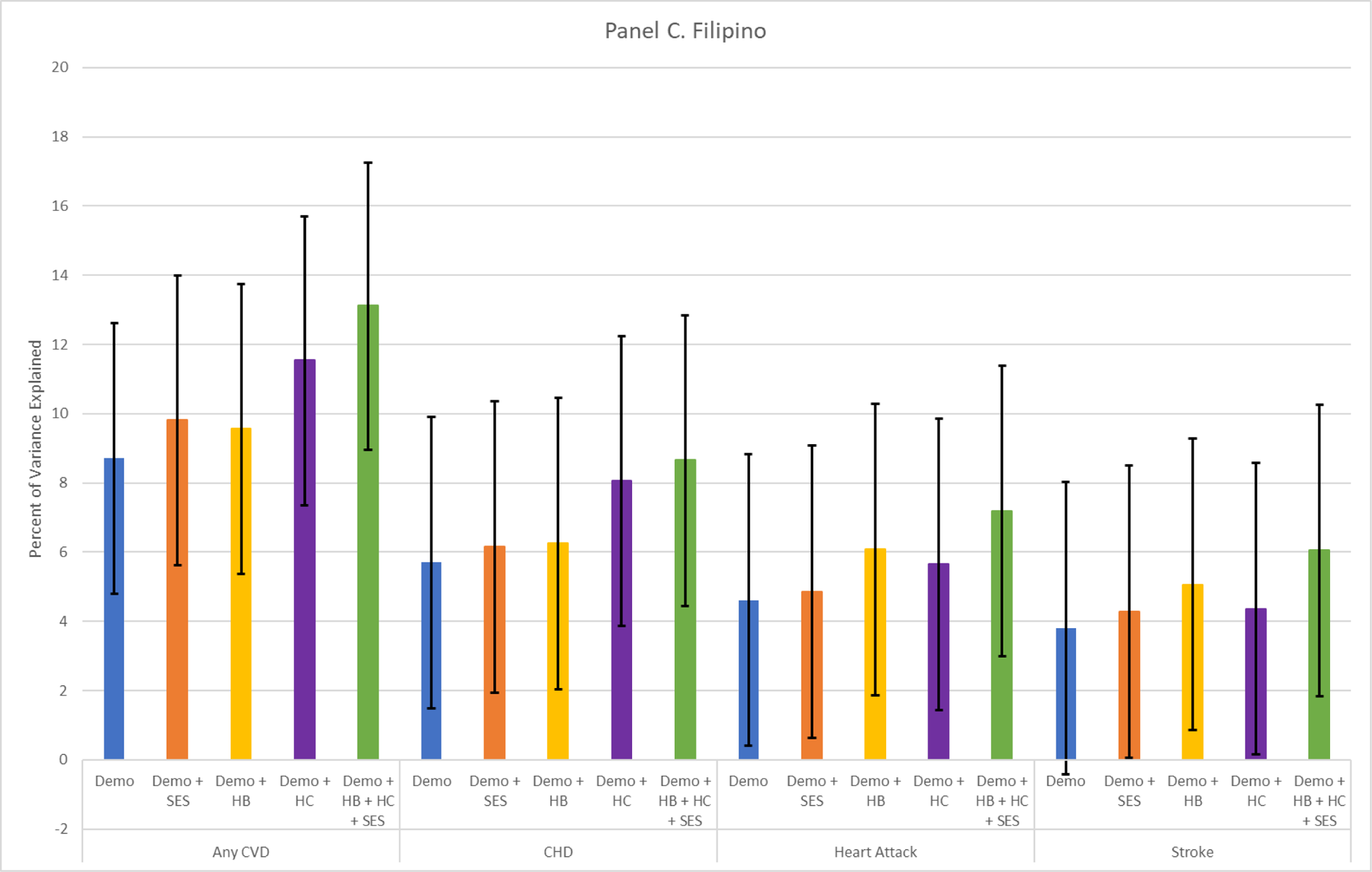
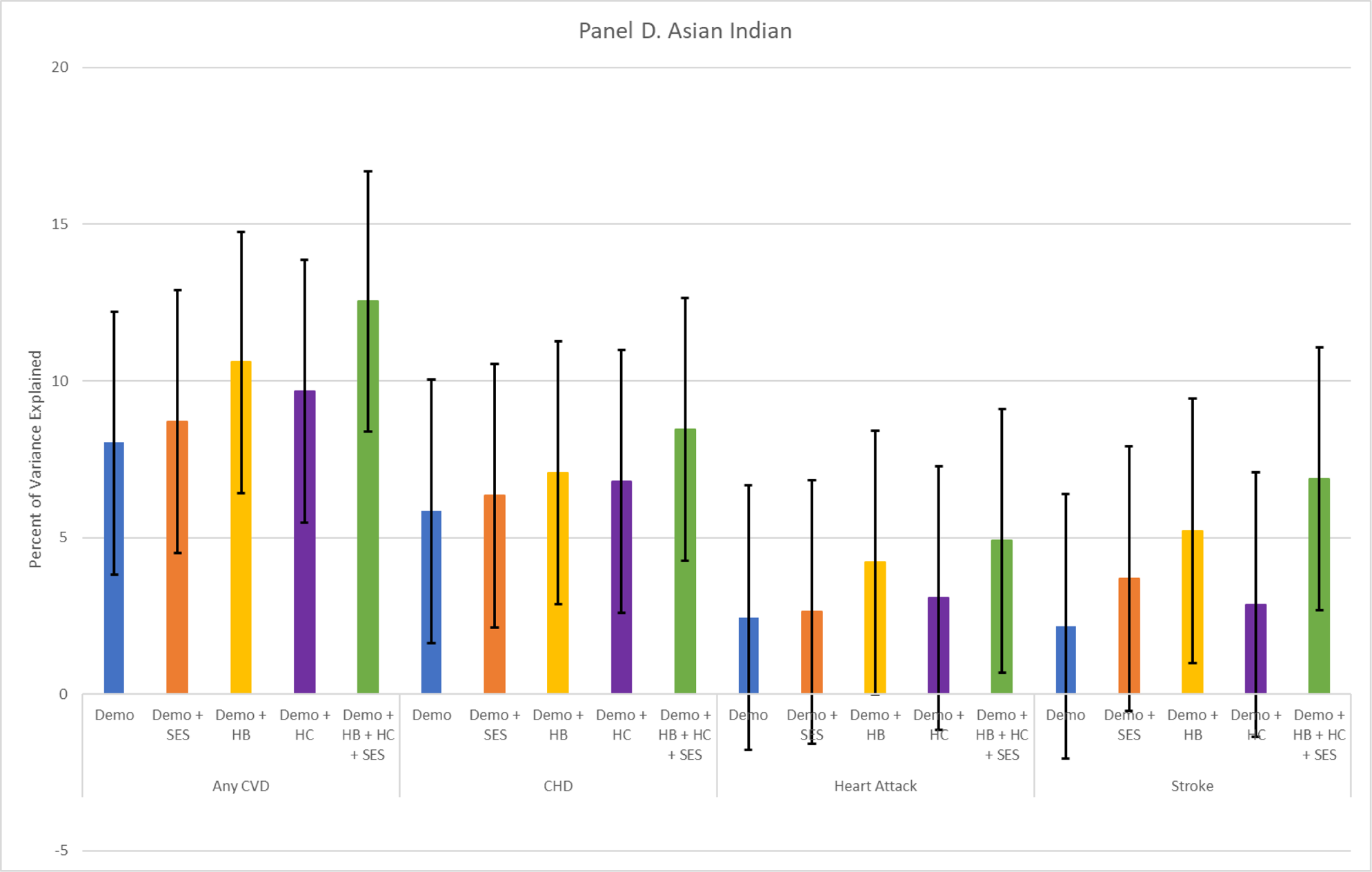
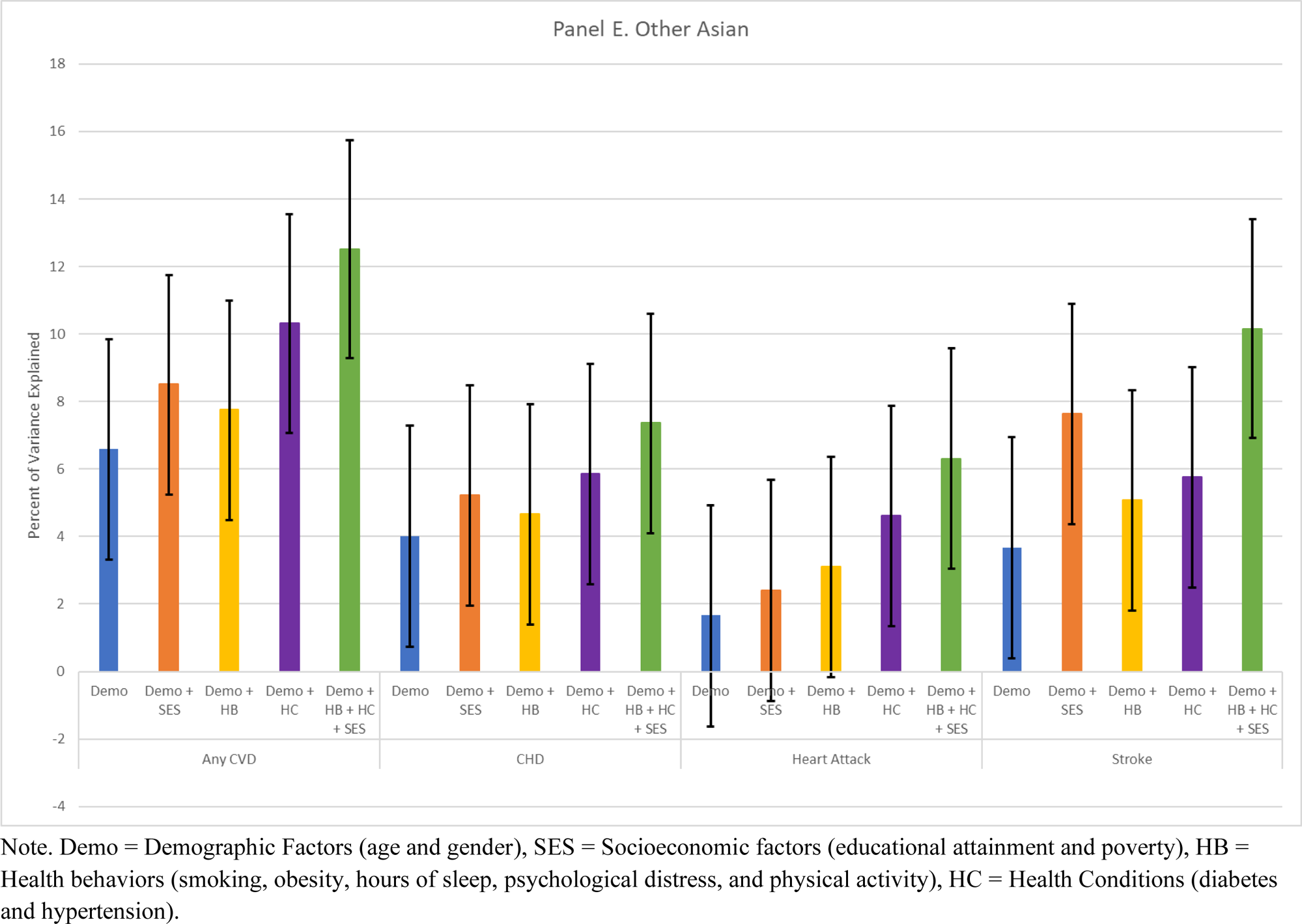
Explained Variance by Race and Outcome

**Table 2.** Percent of Variance Explained by Demographic, Behavioral, Health Factor, and Socioeconomic Factors, All Asian Groups, N = 10,353.

| Model Covariates | R-Squared (95% Confidence Interval) |  |  |  |
| --- | --- | --- | --- | --- |
|  | Any CVD | CHD | Heart Attack | Stroke |
| Demographic | 7.04 (5.12, 8.95) | 4.75 (2.83, 6.67) | 2.71 (0.78, 4.63) | 2.97 (1.04, 4.89) |
| Demographic + SES | 7.63 (5.71, 9.54) | 4.97 (3.05, 6.90) | 2.91 (0.98, 4.83) | 3.84 (1.92, 5.76) |
| Demographic + Health Behavior | 7.87 (5.95, 9.78) | 5.09 (3.17, 7.00) | 3.44 (1.51, 5.36) | 4.00 (2.08, 5.92) |
| Demographic + Health Condition | 10.05 (8.14, 11.95) | 6.57 (4.65, 8.49) | 4.13 (2.21, 6.05) | 4.23 (2.31, 6.15) |
| Demographic + Health Behavior + Health Condition + SES | 11.00 (9.09, 12.90) | 6.86 (4.94, 8.77) | 4.87 (2.95, 6.79) | 5.75 (3.83, 7.67) |

**Table 3.** Percent of Variance Explained by Demographic, Behavioral, Health Factor, and Socioeconomic Factors, Chinese, N = 2,153.

|  | R-Squared (95% Confidence Interval) |  |  |  |
| --- | --- | --- | --- | --- |
| Model Covariates | Any CVD | CHD | Heart Attack | Stroke |
| Demographic | 4.49 (0.27, 8.70) | 3.66 (-0.57, 7.87) | 2.36 (-1.87, 6.58) | 1.44 (-2.79, 5.66) |
| Demographic + SES | 4.95 (0.73, 9.16) | 3.95 (-0.27, 8.16) | 2.54 (-1.69, 6.76) | 1.75 (-2.48, 5.97) |
| Demographic + Health Behavior | 5.54 (1.32, 9.74) | 4.10 (-0.12, 8.31) | 3.07 (-1.16, 7.29) | 3.91 (-0.32, 8.12) |
| Demographic + Health Condition | 7.52 (3.31, 11.71) | 4.91 (0.69, 9.12) | 4.69 (0.47, 8.90) | 3.28 (-0.95, 7.49) |
| Demographic + Health Behavior + Health Condition + SES | 8.82 (4.61, 13.00) | 5.70 (1.48, 9.90) | 5.51 (1.29, 9.71) | 5.89 (1.67, 10.09) |

**Table 4.**
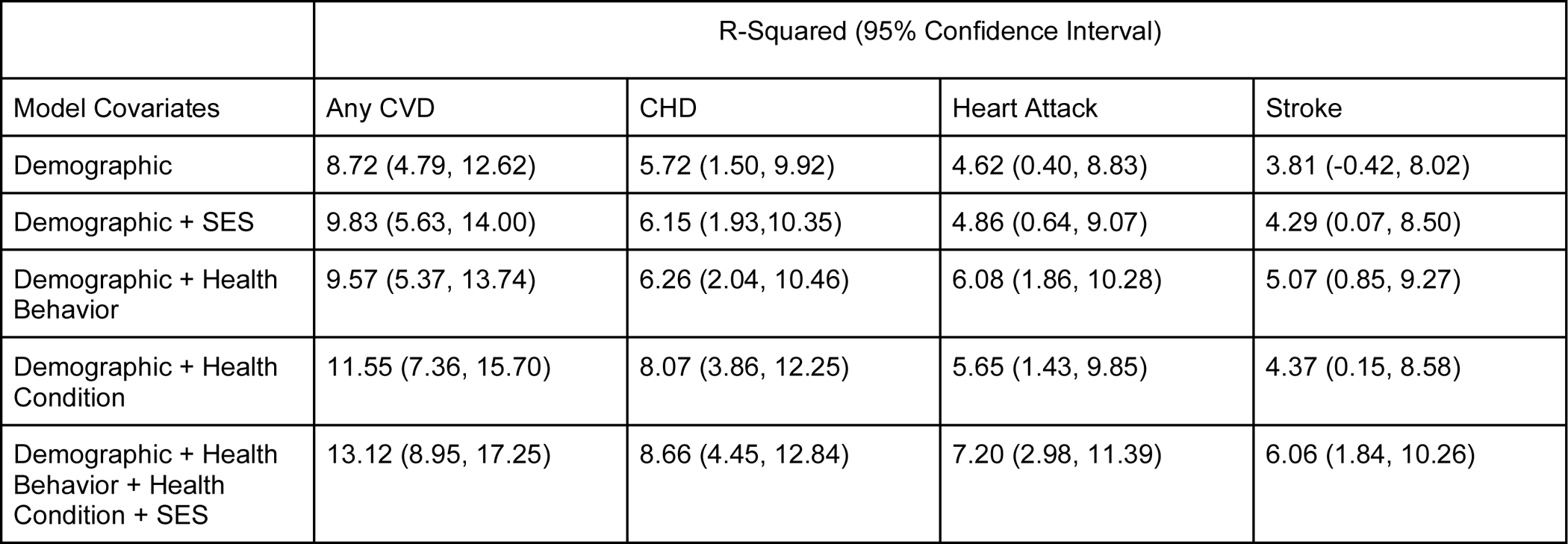
Percent of Variance Explained by Demographic, Behavioral, Health Factor, and Socioeconomic Factors, Filipino, N = 2,466.

**Table 5.**
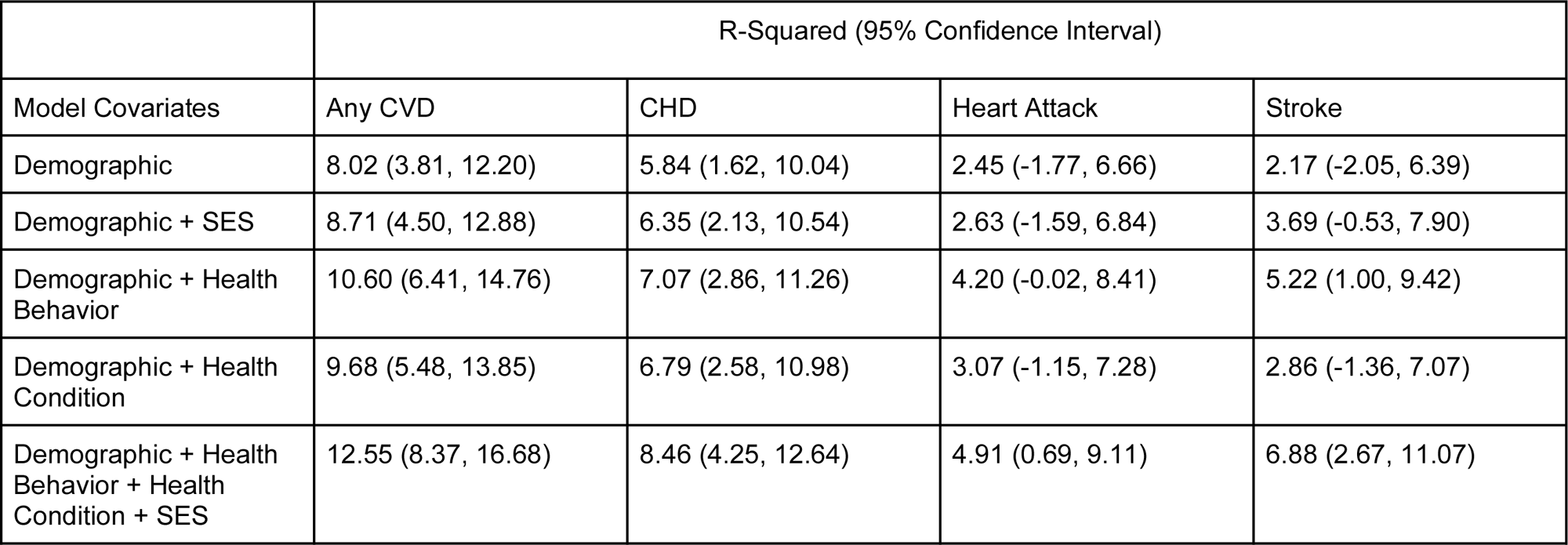
Percent of Variance Explained by Demographic, Behavioral, Health Factor, and Socioeconomic Factors, Asian Indian, N = 2,156.

|  | R-Squared (95% Confidence Interval) |  |  |  |
| --- | --- | --- | --- | --- |
| Model Covariates | Any CVD | CHD | Heart Attack | Stroke |
| Demographic | 8.02 (3.81, 12.20) | 5.84 (1.62, 10.04) | 2.45 (-1.77, 6.66) | 2.17 (-2.05, 6.39) |
| Demographic + SES | 8.71 (4.50, 12.88) | 6.35 (2.13, 10.54) | 2.63 (-1.59, 6.84) | 3.69 (-0.53, 7.90) |
| Demographic + Health Behavior | 10.60 (6.41, 14.76) | 7.07 (2.86, 11.26) | 4.20 (-0.02, 8.41) | 5.22 (1.00, 9.42) |
| Demographic + Health Condition | 9.68 (5.48, 13.85) | 6.79 (2.58, 10.98) | 3.07 (-1.15, 7.28) | 2.86 (-1.36, 7.07) |
| Demographic + Health Behavior + Health Condition + SES | 12.55 (8.37, 16.68) | 8.46 (4.25, 12.64) | 4.91 (0.69, 9.11) | 6.88 (2.67, 11.07) |

**Table 6.**
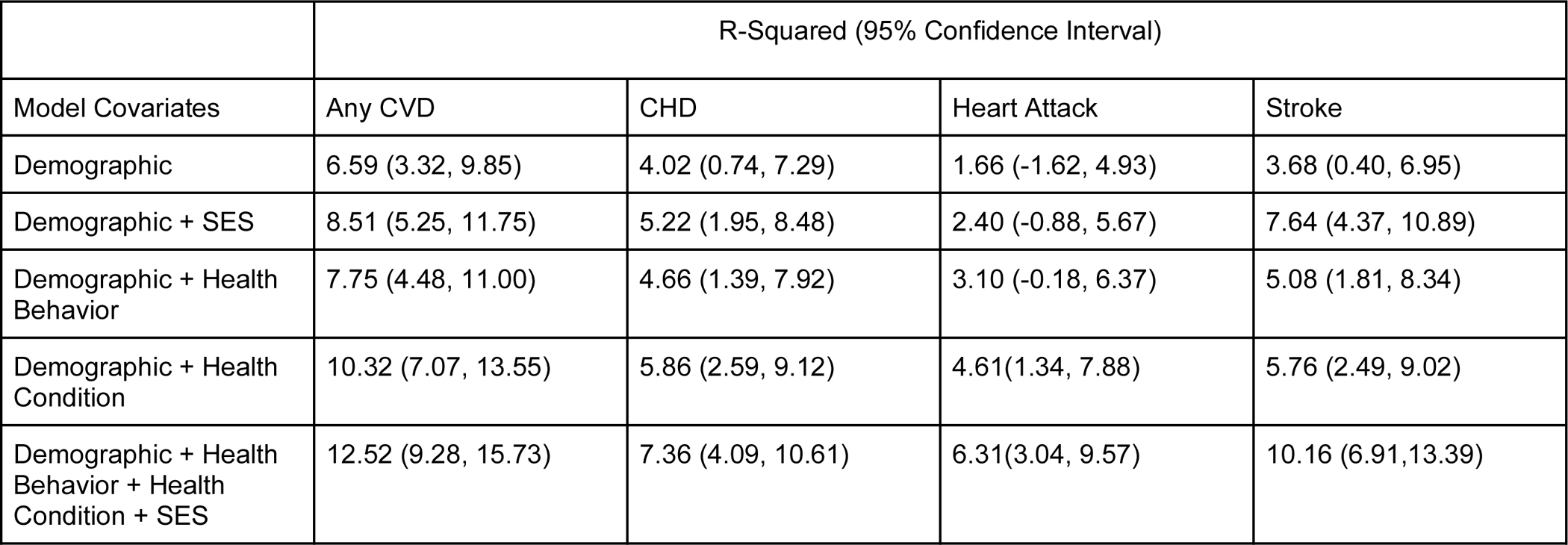
Percent of Variance Explained by Demographic, Behavioral, Health Factor, and Socioeconomic Factors, Other Asian, N = 3,578.

|  | R-Squared (95% Confidence Interval) |  |  |  |
| --- | --- | --- | --- | --- |
| Model Covariates | Any CVD | CHD | Heart Attack | Stroke |
| Demographic | 6.59 (3.32, 9.85) | 4.02 (0.74, 7.29) | 1.66 (-1.62, 4.93) | 3.68 (0.40, 6.95) |
| Demographic + SES | 8.51 (5.25, 11.75) | 5.22 (1.95, 8.48) | 2.40 (-0.88, 5.67) | 7.64 (4.37, 10.89) |
| Demographic + Health Behavior | 7.75 (4.48, 11.00) | 4.66 (1.39, 7.92) | 3.10 (-0.18, 6.37) | 5.08 (1.81, 8.34) |
| Demographic + Health Condition | 10.32 (7.07, 13.55) | 5.86 (2.59, 9.12) | 4.61(1.34, 7.88) | 5.76 (2.49, 9.02) |
| Demographic + Health Behavior + Health Condition + SES | 12.52 (9.28, 15.73) | 7.36 (4.09, 10.61) | 6.31(3.04, 9.57) | 10.16 (6.91,13.39) |

### Additional Variance in CVD Explained by Socioeconomic Characteristics

The additional variance explained by the inclusion of socioeconomic characteristics (SES; i.e., level of educational attainment, and Federal Poverty Level), over and above the model including demographic characteristics as a predictor differed across outcomes and for all AA subgroups (orange bars, Figure 1, Panels A-E). Among all AA participants, SES characteristics consistently contributed the least amount of additional explained variance, such that estimates for R-squared closely resembled the estimates from models only including demographic characteristics ranging from 2.9% heart attack to 7.6% for any CVD. Similarly, SES characteristics also consistently contributed the least amount of explained variance for Chinese participants (at most an additional 0.46%), such that estimates ranged from 1.8% stroke to 5.0% any CVD. For Filipino participants, SES characteristics added an additional 1.1% of variance that can be explained for any CVD, but for all other CVD events, SES characteristics at most contributed an additional 0.48%, such that estimates ranged from 4.3% stroke to 9.8% any CVD. Among Asian Indian participants, SES characteristics at most contributed 1.5% additional variance that can be explained, such that estimates ranged from 2.6% heart attack to 8.7% any CVD. Among Other Asian participants, SES characteristics contributed up to 4% of additional explained variance, such that estimates ranged from 2.4% heart attack to 8.5% any CVD. SES characteristics more than doubled the explained variance for stroke beyond models with demographic characteristics only (7.6% versus 3.7% respectfully) amongst Other Asian participants.

### Additional Variance in CVD Explained by Health Behaviors

The additional variance explained by including health behaviors over and above the model with only demographic characteristics as a predictor differed across outcomes and for all AA subgroups (yellow bars, Figure 1, Panels A-E). Among all AA participants, health behaviors contributed at most an additional 1.0% of explained variance, such that estimates ranged from 3.4% heart attack to 7.9% any CVD. Among Chinese participants, health behaviors only contributed about 0.5% of additional explained variance for any CVD, CHD, and heart attack, such that estimates ranged from 3.1% for heart attack to 5.5% for any CVD. Of note, health behaviors did contribute an additional 2.5% to stroke, explaining 3.9% of variance to the outcome of stroke compared to 1.4% of variance explained in models with only demographic characteristics. Among Filipino participants, health behaviors contributed at most 1.5% of additional explained variance beyond the baseline model, such that estimates ranged from 5.1% stroke to 9.6% any CVD. Among Asian Indian participants, health behaviors consistently contributed at least 2.0% of the additional explained variance for any CVD, CHD, and heart attack, but an additional 3.1% to stroke; estimates ranged from 4.2% heart attack to 10.6% for any CVD. Among Other Asian participants, health behaviors contributed at most 1.4% additional explained variance, such that estimates ranged from 0.4% CHD to 1.4% heart attack and stroke.

### Additional Variance in CVD Explained by Health Conditions

The additional variance explained by including health conditions over and above the model with only demographic characteristics as a predictor differed across outcomes and for all AA ethnic groups (Figure 1, Panels A-E). For all AA participants, health conditions contributed at least 1.4% additional explained variance beyond demographics such that estimates ranged from 4.1% (heart attack) to 10.1% any CVD. Among Chinese participants, health conditions contributed at least 1.8% of additional explained variance such that estimates ranged from 3.3% stroke to 7.5% any CVD. Among Filipino participants, health conditions contributed at least 0.5% additional explained variance such that estimates ranged from 4.4% stroke to 11.6% any CVD. Among Asian Indian participants, health conditions contributed at least 0.6% of the additional explained variance such that estimates ranged from 2.9% stroke to 9.7% amu CVD. Among Other Asian participants, health conditions consistently contributed at least 1.8% of additional explained variance across outcomes, such that estimates ranged from 4.6% heart attack to 10.3% any CVD.

### Additional Variance in CVD Explained by Socioeconomic Characteristics, Health Behaviors, and Health Conditions

When including socioeconomic characteristics and both sets of CVD predictors–health behaviors and health conditions (green bars in Figure 1, Panels A-E)--the additional variance explained increased relative to the variance explained by demographic characteristics alone for all AA subgroups. Among all AA participants, estimates ranged from 4.9% heart attack to 11% any CVD. For Chinese participants, estimates ranged from 5.5% to 8.8% any CVD. For Filipino participants, estimates ranged from 6.1% stroke to 13.1% any CVD. For Asian Indian participants, estimates ranged from 4.9% heart attack to 12.6% any CVD. For Other Asian participants, estimates ranged from 6.3% heart attack to 12.5% any CVD.

## DISCUSSION

Results from the present study show that the variance explained by sociocultural and behavioral factors across a range of CVD outcomes and risk factors for disaggregated AA ethnic groups show considerable heterogeneity corroborated previous research.^13^ A greater relative burden of CVD was observed in Filipino Americans, particularly for CHD, heart attack, and any CVD, while Other Asians had a greater burden of stroke. Results also indicate that CVD-related health behaviors and health conditions may increase or decrease the prevalence of CVD risk factors across AA ethnic groups. Again, Filipino Americans are more likely to engage in health behaviors known to be linked to CVD, such as smoking tobacco, being physically inactive, and sleeping less than other AA ethnic groups. Filipino Americans are also more likely to report CVD-related health conditions such as diabetes and hypertension. However, health behaviors explained a greater amount of variance only for Asian Indians, while health conditions explained a greater amount of variance for all ethnic groups except for Asian Indians. Collectively, study findings emphasize the heterogeneity of CVD trends in AA ethnic groups, amplifying the need for tailored prevention and intervention efforts for the different ethnic groups.

For all AA participants, demographic characteristics (age and gender) alone explained at least half of the variance from combined predictors. However, the contribution of explained variance due to demographics seems especially pronounced for Filipino participants, as shown by having the highest percentage of explained variance across all CVD outcomes between the ethnic groups and when compared to the entire AA sample. This finding suggests that across all CVD outcomes, Filipinos, particularly older Filipino males, should be targeted for CVH promotion, corroborating previous research.^24^ Contributions of explained variance due to SES characteristics also differed across groups, with the largest contributions found among the Other Asian group across all CVD outcomes when compared to the other ethnic groups, and when compared to the entire AA sample. As aforementioned, the publicly available NHIS dataset does not disaggregate the Other Asian group; as such, we are unable to unpack these findings fully. However, previous research shows that educational attainment among Korean Americans (a possible population included within the Other Asian group) was associated with hypertension; less education was significantly and negatively correlated with risk for hypertension.^25^ The Other Asian group does have a larger percentage (31%) of participants with only a high school degree or less than a high school education. Based on prior research,^26^ we postulate that acculturation may explain the larger contribution of explained variance of SES among the Other Asian group such that they have a more recent immigration history to the U.S.,^27^ thus contributing to their smaller sample sizes and necessary aggregation. This suggests that less time spent in the U.S. means less time to establish themselves to attain higher levels of education and secure better paying jobs.

Differences in health behaviors and conditions were also significant predictors of CVD between different AA ethnic groups. Compared to Chinese, Filipino, and Other Asians, health behaviors were an essential predictor for all CVD outcomes among Asian Indians. Inversely, existing health conditions were significant predictors of CVD for all AA ethnic groups compared to Asian Indians. Such differences underscore the complexity of which factors are critical for different AA ethnic groups that situate them at a higher burden for CVD. Our findings support previous research demonstrating heterogeneity in CVD burden among AA ethnic groups.^9, 28^ Indeed, a recently published study analyzing the same NHIS data disaggregated by AA ethnic groups showed significant increases and decreases in the prevalence of various CVD-related risk factors across AA ethnic groups.^13^

When combining all predictors, Filipinos had the highest percentage of variance explained for any CVD, CHD, and heart attack. Our findings support prior limited research, which shows that Filipinos have a significantly disproportionate burden of CVD when compared to other racial/ethnic groups, including non-Hispanic whites.^7^ Despite these alarming rates and the fact that Filipinos constitute the third largest AA ethnic group, they remain disproportionately understudied.^29^ Our results indicate that we must continue to engage Filipinos in research to improve prevention and intervention strategies such as risk factor detection, health literacy, and disease management.^30^ Only stroke for the Other Asian group had a higher percentage of variance explained with almost twice the amount of explained variance for stroke than the aggregate AA group and almost 4% more explained variance than each of the other subgroups. This finding underscores the importance of also differentiating between cardiovascular and cerebrovascular disease outcomes among AAs, particularly since there is heterogeneity in cardiovascular and cerebrovascular mortality, with Vietnamese women (a group potentially included in the Other Asian category) having the highest age-standardized mortality rate due to cerebrovascular disease (e.g., stroke) when compared to other AA ethnic groups.^31^

The predominant strength of our analysis is the exposition of detailed contemporary national prevalence statistics for leading causes of CVD outcomes and risk factors in disaggregated AA ethnic groups. There are several limitations to be noted. First, the analysis was restricted to a sample of Chinese, Asian Indian, and Filipino participants and an aggregated “Other Asian” category due to the small sample size for other Asian subgroups in the NHIS. However, there are very few data sets other than the NHIS in the U.S. that disaggregate data on AAs to include details on demographics, socioeconomic, and CVD risk factors. Thus, this study contributes to increased understanding of how to address CVD in priority populations. Additionally, this data is cross-sectional. Readers should be cautioned about determining causal relationships between demographic, socioeconomic, and CVD-related health behaviors and conditions, even though predictive modeling is often a first step in understanding these causal relationships. Moreover, this data is self-reported.

In addition, findings from this study explained, at most, only 13% of variance. However, this percentage is similar to previous research, such as Hamad et al.,^19^ which found 14% variance that can be explained by demographic, socioeconomic, and genetic predictors. Additionally, like Hamad et al.,^19^ the modest percentage of explained variance we found reflects the possible role of construct measurement error, randomness, and other unmeasured factors. For example, a systematic review on social determinants of cardiometabolic diseases among AA ethnic groups found that acculturation, social context, and health literacy may explain heterogeneity of CVD risk and outcomes.^32^ As such, future research should continue to look at other CVD-related factors, including factors beyond the individual level, such as discrimination per the NIHMD Research Framework. Data shows that AA experienced increased race-based discrimination during the COVID-19 pandemic.^33^ This spike in documented anti-AA discrimination has been correlated with various adverse health outcomes.^34^ Although CVD-related deaths did increase for AAs during the U.S. COVID-19 pandemic in 2020 compared with a historical control (2019), studies linking anti-AA discrimination with increased CVD remain limited.

## CONCLUSION

Study results confirm previous research demonstrating heterogenous trends in CVD among AAs, particularly that factors that help explain CVD operate differently across AA ethnic groups. This study contributes to addressing CVD disparities and promoting health equity^35–37^ by conducting research that is intentional about data disaggregation, appropriately representing AA ethnic groups. Additionally, although findings signal worrisome trends, healthcare providers, researchers, and policymakers have immense opportunities to alleviate the burden of CVD in AA populations by developing patient-centered strategies that promote continued engagement in CVD screening, heart-healthy behaviors, and CVD disease management. Our study’s findings can inform such targeted efforts, particularly the development of culturally-responsive interventions to address differing CVD risk factors and improve CVD outcomes among specific AAs ethnic groups.

## Data Availability

We used secondary data from the 2013-2018 National Health Interview Survey (NHIS) obtained through the Integrated Public Use Microdata Series (IPUMS). The NHIS is a nationally representative health survey and a primary federal source of health information. Detailed information on the survey design and methods can be found elsewhere (http://www.cdc.gov/nchs/nhis.htm).

## Sources of Funding

Dr. Adrian Bacong received funding from the Stanford Cardiovascular Institute Seed Grant.

## Disclosures

None of the authors have any disclosures to report.

## Notes

### Competing Interest Statement

The authors have declared no competing interest.

### Author Declarations

This is a secondary data analysis of publicly available data. No IRB approval was needed.

## References

1. Budiman A, Ruiz NG. Asian Americans are the fastest-growing racial or ethnic group in the U.S. The Pew Research Center. 2021.

2. US Census Bureau. 2013-2017 American Community Survey 5-year estimates [Internet]. Available from: https://www.census.gov/programs-surveys/acs/technical-documentation/table-and-geography-changes/2017/5-year.html. Accessed 2024 Mar 19.

3. Shetty NS, Patel N, Gaonkar M, Kalra R, Li P, Pavela G, Arora G, Arora P. Trends of cardiovascular health in Asian American individuals: A national health and nutrition examination survey study. Am J Prev Cardiol. 2023 May 26:100509.

4. Kwan TW, Wong SS, Hong Y, Kanaya AM, Khan SS, Hayman LL, Shah SH, Welty FK, Deedwania PC, Khaliq A, Palaniappan LP. Epidemiology of diabetes and atherosclerotic cardiovascular disease among Asian American adults: implications, management, and future directions: a scientific statement from the American Heart Association. Circulation. 2023 May 8.

5. Gordon NP, Lin TY, Rau J, Lo JC. Aggregation of Asian-American subgroups masks meaningful differences in health and health risks among Asian ethnicities: an electronic health record-based cohort study. BMC Public Health. 2019 Dec;19(1):1–4.

6. Kianoush S, Rifai MA, Jain V, Samad Z, Rana J, Dodani S, et al. Prevalence and Predictors of Premature Coronary Heart Disease Among Asians in the United States: A National Health Interview Survey Study. Current Problems in Cardiology. 2023 Jul;48(7):101152.

7. Rivera FB, Cha SW, Ansay MFM, Taliño MKV, Flores GP, Nguyen RT, Bonuel N, Happy Araneta MR, Volgman AS, Shah N, Vahidy F, Cainzos-Achirica M. Cardiovascular disease in Filipino American men and women: a 2023 update. Am Heart J. 2023;266:1–13. doi: 10.1016/j.ahj.2023.07.015.

8. Shah NS, Luncheon C, Kandula NR, Khan SS, Pan L, Gillespie C, et al. Heterogeneity in Obesity Prevalence Among Asian American Adults. Ann Intern Med. 2022 Nov;175(11):1493– 500.

9. Kianoush S, Al Rifai M, Merchant AT, Jia X, Samad Z, Bhalla A, et al. Heterogeneity in the prevalence of premature hypertension among Asian American populations compared with white individuals: A National Health Interview Survey study. International Journal of Cardiology Cardiovascular Risk and Prevention. 2022 Sep;14:200147.

10. Nandagiri V, Vannemreddy S, Spector A. Sleep disparities in Asian Americans: a comprehensive review. Journal of Clinical Sleep Medicine. 2023 Feb;19(2):393–402.

11. Agaku IT, Odani S, Okuyemi KS, Armour B. Disparities in current cigarette smoking among US adults, 2002–2016. Tob Control. 2019 May 30;tobaccocontrol-2019-054948.

12. Nguyen AB. Disaggregating Asian American and Native Hawaiian and Other Pacific Islander (AANHOPI) Adult Tobacco Use: Findings from Wave 1 of the Population Assessment of Tobacco and Health (PATH) Study, 2013–2014. J Racial and Ethnic Health Disparities. 2019 Apr 15;6(2):356–63.

13. Lim A, Elias S, Benjasirisan C, Byiringiro S, Chen Y, Turkson-Ocran R, et al. Heterogeneity in the Prevalence of Cardiovascular Risk Factors by Ethnicity and Birthplace Among Asian Subgroups: Evidence From the 2010 to 2018 National Health Interview Survey. JAHA. 2024 Mar 5;13(5):e031886.

14. Huang YC, Garcia AA. Culturally-tailored interventions for chronic disease self-management among Chinese Americans: a systematic review. Ethn Health. 2020;25(3):465–484. doi: 10.1080/13557858.2018.1432752.

15. Joo JY. Effectiveness of culturally tailored diabetes interventions for Asian immigrants to the United States: a systematic review. The Diabetes Educator. 2014 Sep;40(5):605–15.

16. National Institute on Minority Health and Health Disparities (2017). NIMHD Research Framework.

17. Kessler RC, Andrews G, Colpe LJ, Hiripi E, Mroczek DK, Normand SL, Walters EE, Zaslavsky AM. Short screening scales to monitor population prevalences and trends in non-specific psychological distress. Psychological medicine. 2002 Aug;32(6):959–76.

18. Craig CL, Marshall AL, Sjöström M, Bauman AE, Booth ML, Ainsworth BE, Pratt M, Ekelund UL, Yngve A, Sallis JF, Oja P. International physical activity questionnaire: 12-country reliability and validity. Medicine & science in sports & exercise. 2003 Aug 1;35(8):1381–95.

19. Hamad R, Glymour MM, Calmasini C, Nguyen TT, Walter S, Rehkopf DH. Explaining the variance in cardiovascular disease risk factors: a comparison of demographic, socioeconomic, and genetic predictors. Epidemiology. 2022 Jan 1;33(1):25–33.

20. Menard S. Coefficients of determination for multiple logistic regression analysis. The American Statistician. 2000 Feb 1;54(1):17–24.

21. Mittlböck M, Schemper M. Explained variation for logistic regression. Statistics in medicine. 1996 Oct 15;15(19):1987–97.

22. Cohen, Jacob, Patricia Cohen, Stephen G. West, and Leona S. Aiken. Applied multiple regression/correlation analysis for the behavioral sciences. Routledge, 2013.

23. StataCorp. 2021. Stata Statistical Software: Release 17. College Station, TX: StataCorp LLC.

24. Satish P, Sadaf MI, Valero-Elizondo J, Grandhi GR, Yahya T, Zawahir H, Javed Z, Mszar R, Hanif B, Kalra A, Virani S. Heterogeneity in cardio-metabolic risk factors and atherosclerotic cardiovascular disease among Asian groups in the United States. American Journal of Preventive Cardiology. 2021 Sep 1;7:100219.

25. Kim MT, Kim KB, Juon HS, Hill MN. Prevalence and factors associated with high blood pressure in Korean Americans. Ethnicity & Disease. 2000 Oct 1;10(3):364–74.

26. Yuemeng LI, Alicia ZH, Austin LE, Singh J, Palaniappan LP, Srinivasan M, Elfassy, Valero-Elizondo J, Eugene YA. Association of acculturation with cardiovascular risk factors in Asian-American subgroups. American journal of preventive cardiology. 2023 Mar 1;13:100437.

27. Guadamuz JS, Kapoor K, Lazo M, Eleazar A, Yahya T, Kanaya AM, Cainzos-Achirica M, Bilal U. Understanding immigration as a social determinant of health: cardiovascular disease in Hispanics/Latinos and South Asians in the United States. Current atherosclerosis reports. 2021 Jun;23:1–2.

28. Ramaswamy P, Mathew Joseph N, Wang J. Health Beliefs Regarding Cardiovascular Disease Risk and Risk Reduction in South Asian Immigrants: An Integrative Review. J Transcult Nurs. 2020 Jan;31(1):76–86.

29. Arigorat EJ, Begonia K, Franklin M, Honsky J. Assessment of Electronic Health Literacy Among Filipino Americans. CIN: Computers, Informatics, Nursing. 2024 Jul 1;42(7):530–6.

30. Abesamis CJ, Fruh S, Hall H, Lemley T, Zlomke KR. Cardiovascular health of Filipinos in the United States: a review of the literature. J Transcult Nurs. 2016;27(5):518–28.

31. Shah, N. S., Xi, K., Kapphahn, K. I., Srinivasan, M., Au, T., Sathye, V., … & Palaniappan, L. P. (2022). Cardiovascular and cerebrovascular disease mortality in Asian American subgroups. Circulation: cardiovascular quality and outcomes, 15(5), e008651.

32. Min LY, Islam RB, Gandrakota N, Shah MK. The social determinants of health associated with cardiometabolic diseases among Asian American subgroups: a systematic review. BMC Health Services Research. 2022 Feb 25;22(1):257.

33. Stop AAPI Hate. (2023). Taxonomy report: Key findings. Stop AAPI Hate. https://stopaapihate.org/wp-content/uploads/2023/10/23-SAH-TaxonomyReport-KeyFindings-F.pdf

34. Wu C, Qian Y, Wilkes R. Anti-Asian discrimination and the Asian-white mental health gap during COVID-19. In Race and Ethnicity in Pandemic Times 2021 Sep 30 (pp. 101–117). Routledge.

35. Angell SY, McConnell MV, Anderson CA, Bibbins-Domingo K, Boyle DS, Capewell S, Ezzati M, De Ferranti S, Gaskin DJ, Goetzel RZ, Huffman MD. The American Heart Association 2030 impact goal: a presidential advisory from the American Heart Association. Circulation. 2020 Mar 3;141(9):e120–38.

36. Churchwell K, Elkind MS, Benjamin RM, Carson AP, Chang EK, Lawrence W, Mills A, Odom TM, Rodriguez CJ, Rodriguez F, Sanchez E. Call to action: structural racism as a fundamental driver of health disparities: a presidential advisory from the American Heart Association. Circulation. 2020 Dec 15;142(24):e454–68.

37. Lloyd-Jones DM, Elkind M, Albert MA. American Heart Association’s 2024 impact goal: every person deserves the opportunity for a full, healthy life. Circulation. 2021 Nov 2;144(18):e277–9.

